# A Retrospective Cohort Analysis Investigating Determinants and Rates of Retention among Children and Adolescents Receiving Antiretroviral Therapy in Center and East Regions of Cameroon

**DOI:** 10.1101/2025.08.07.25333255

**Authors:** Bryan Shaw, Nkenkeu Kieuvo Ignace, Mbogwe Ndimumeh, Lauren Mathae, Ryan Neil, Chibuike Mark Ogbuabo, Julienne Mesumbe, Ignace Gashongore, Deus Bazira

**Affiliations:** Center for Global Health Practice and Impact USA, Georgetown University, Washington, D.C., USA; Center for Global Health Practice and Impact Cameroon, Yaoundé, Cameroon

## Abstract

**Background:** Children and adolescents (0 – 15 years) living with HIV (ACLHIV) in low-resource settings face individual, household, and systemic challenges to engagement and retention in HIV care and antiretroviral treatment (ART) adherence. As a result, ACLHIV register poorer care and treatment outcomes. We describe care retention outcomes and factors that influence them among ACLHIV enrolled in ART in Cameroon.

**Methods:** We explored trends in two-year follow-up and sociodemographic and clinical factors associated with retention in care among ACLHIV living in the Center and East Regions. We retrospectively analyzed anonymized medical records (*n* = 1,319) from a cohort of all ACLHIV initiating ART for a three-year period between October 1, 2019, to September 30, 2022. Data were obtained from electronic medical records from 66 HIV treatment sites. Trends in retention in care among ACLHIV on ART were described using Kaplan-Meier survival estimates. Cox proportional analysis was performed to identify factors associated with retention in care.

**Results:** Retention in care among ACLHIV in both regions at months 6, 12, 18, and 24 was 86.2%, 82.6%, 79.8%, and 77.2%, respectively. After 24 months, retention was higher in the Center Region (83.1%) compared to the East Region (71.5%). In adjusted regression models, female ACLHIV were more likely to be retained in care in the Center Region but less likely in the East Region compared to boys after 24 months. Clinical treatment factors, such as current use of a dolutegravir and receipt of ART through differentiated service delivery models, were associated with a considerably reduced risk of loss to follow-up. Unexpectedly, we found that very young children (<5 years) were less likely to be retained in care than older adolescents (5-15 years), which could be partially explained by local context of care.

**Conclusion:** This study suggests that Cameroon is lagging behind targets for HIV retention in care for ACLHIV. More effective and targeted clinical and social behavioral interventions are needed to address barriers to long term engagement in care by ACLHIV. Targeted research studies and improved data-driven, process monitoring is needed to determine other underlying factors that lead to sub-optimal care retention outcomes among this sub-population.

## INTRODUCTION

Retention in care is a critical aspect of the global response to the HIV/AIDS epidemic, encompassing the engagement of HIV-positive individuals in ongoing medical care from point of diagnosis through initiation and lifelong adherence to antiretroviral therapy (ART) ^(1,2)^. With the adoption of the World Health Organization’s (WHO) 2015 ’HIV Treat All’ strategy, it is increasingly imperative to optimize long-term retention in treatment for all populations, including children and adolescents (0-15 years) living with HIV (ACLHIV) ^(3,4)^. In 2020, UNICEF estimated that there were more than 2.5 million ACLHIV in sub-Saharan Africa (SSA), and adolescents and young people remain at high risk for new HIV infections. ACLHIV, in particular, are at increased risk of AIDS-related mortality, experience lower rates of retention in HIV care and treatment, and have sub-optimal HIV virologic suppression rates compared to adults across all income settings ^(3,5,6)^. The growing number of ACLHIV is driven by continued increases in HIV incidence through horizontal transmission, particularly among adolescent girls, and by the success of pediatric ART programs in increasing survival of children with perinatally-acquired HIV ^(7)^.

Low- and middle-income countries (LMIC) face unique challenges in supporting a large and growing population of youth requiring ART, but these efforts are often undermined by continuing interruptions in treatment within this age group ^(8,9)^. Studies indicate that ACLHIV in LMIC typically have higher attrition rates compared to adults, ranging from 25% to 35% in a series of ACLHIV cohorts in SSA. This underscores the urgent need for effective interventions targeting retention ^(10,11,12)^. Several retention strategies for ACLHIV have been implemented in LMIC, in contrast to high-income settings, including treatment decentralization, task-shifting to expand the pool of healthcare providers authorized to prescribe and dispense ART, and differentiated care models to improve ART access and enhance patient experience. These strategies are showing promise in LMIC ^(6,13–15)^; however, socio-structural challenges, including limited healthcare resources, high transportation costs, and socio-economic factors, continue to negatively impact patient retention ^(15)^. Moreover, evidence-based interventions specifically focusing on children and adolescents are very scarce in LMIC ^(5–6,13)^.

A study by Nkefou et al. (2024) ^(16)^ shows that despite steady progress in the prevention of mother-to-child transmission in Cameroon, transmission rates and the number of children diagnosed with HIV remain stubbornly high. In 2021, an estimated 33,000 [24,000 – 38,000] ACLHIV were living in Cameroon ^(17)^, representing 6.6% of all Cameroonians living with HIV (PLHIV). Although targeted interventions have reduced new infections among this population of ACLHIV, Cameroon remains one of 10 countries accounting for 75% of new pediatric HIV infections ^(18)^. HIV continues to be a leading cause of under-five and child mortality in the country ^(19)^, and ART coverage among children remains very low compared to that of Cameroonian adults ^(20)^. Mother-to-child HIV transmission is estimated at 7% nationally, with significant geographical variation, reaching up to 17.5% in the Adamaoua Region ^(21)^.

As Cameroon advances towards its targets, there is a critical need for interventions specifically designed to improve retention of ACLHIV in ART treatment and care. This paper explores trends in two-year follow-up and analyzes sociodemographic and clinical factors associated with retention in care among ACLHIV in the Center and East Regions of Cameroon.

## MATERIALS & METHODS

### Study design

This study was a retrospective cohort design utilizing quantitative data collection and analysis. Data was extracted from the cohort on 10/01/2024 covering the study period from 10/01/2019 to 09/30/2022. It focused on ACLHIV (0 – 15 years) who were receiving ART across 66 public and private HIV treatment sites in the Center (*n* = 47) and East Regions (*n* = 19) of Cameroon.

Collectively, these treatment sites represent 24 of 30 health districts in the Center Region and all 14 health districts in the East Region, minimizing the effect of group clustering. Selected facilities provide comprehensive healthcare services, including prevention and treatment of pediatric HIV, serving rural, semi-urban, and urban populations. Approximately 40% of all PLHIV in the country live in the two regions.

### Study setting & participants

We abstracted medical records for children and adolescents who initiated ART between October 1, 2019 and September 30, 2022, using patient registers and electronic medical records (EMR). The primary outcome of interest was retention in HIV care and ART adherence.

Sociodemographic and clinical factors were extracted, and each pediatric patient was followed for 24 months to assess their HIV treatment outcomes. This study included HIV-positive children and adolescents (ages 0 - 15) at the time of ART initiation. Only HIV-positive children and adolescents (currently 1 – 20 years of age at the time of study enrollment) with complete information in their records were included, while patients with incomplete information were excluded from the study.

### Sample size determination & sampling

We retrospectively analyzed medical records (*n* = 1,319) from a cohort of ACLHIV (0 – 15 years) who initiated ART during the study period under the Translating Data and Evidence into Impact (TIDE) program, implemented by Georgetown University and funded through U.S. Centers for Disease Control and Prevention (CDC) by the U.S President’s Plan for AIDS Relief (PEPFAR). The health facilities included public, private, faith-based, and other not-for-profit sites. All children and adolescents who initiated care at one of these facilities and met the inclusion criteria were followed for 24 months to assess treatment outcomes, including retention in ART.

### Data collection & variables

The primary outcome of this study was retention in care and on ART adherence for ACLHIV. A patient was considered retained in care if they did not miss an expected clinical contact by more than 28 days after their last completed contact, adhered to their ART regimen, and were not known to have transferred in or out of the facility or died before the end of each 6-month time point within the overall 24-month study period. Research assistants assessed retention in care at 6-, 12-, 18-, and 24-month follow-ups after ART initiation. Retention in care was monitored in accordance with national guidelines, including the implementation of differentiated service delivery, such as multi-month ART dispensing for up to 6 months. Patients who had not experienced any medical events requiring a health facility visit or clinical contact between scheduled routine appointments were not required to return for a provider visit until their next scheduled appointment.

We extracted data from the medical records of children and adolescents using both printed patient records and electronic patient management systems: OpenMRS for the East Region, and the Data Manager (DAMA) system for the Center Region. These data systems are used to monitor HIV care processes and patient outcomes. A data extraction checklist was developed to obtain sociodemographic, clinical, and treatment-related variables. Data was extracted at six-month intervals.

Sociodemographic variables extracted for ACLHIV included: sex, age, height, weight, education level, orphan status, and household size. Additionally, we extracted sociodemographic variables for primary caregivers of the ACLHIV, including: relationship to child, sex, age, marital status, education level, occupation, religion, HIV status, and ART status. Baseline clinical and treatment-related data collected included: recent height and weight, time from HIV-positive test to ART initiation, HIV testing entry point (i.e., testing done at community level or within a health facility), ART regimen at initiation and current regiment if changed after baseline (i.e., dolutegravir (DTG) vs. non-DTG-based regimen), ART dispensation model (i.e., direct service delivery (DSD) vs. non-DSD model), documented opportunistic infections diagnosis, WHO stage of disease, and viral load test results. In this context, DSD models included home-based, family-based, or community-based organization (CBO) dispensing, support group or peer pick-up, and multi-month dispensing at the facility level. Finally, our data extraction checklist included facility-level predictors such as: region of facility, urbanicity, type of facility (i.e., public, private, etc.), and facility tiers (categorized based on increasing patient volume).

However, not all variables were used in analysis due to an unacceptable degree of missingness in certain variables. Data were collected by trained, field-based program Data Management

Associates, who triangulated data from individual files, ART registers, and the electronic databases to create a complete longitudinal record for each case of interest. These Data Management Associates are based at health facilities and tasked with a variety of tasks, including: data extraction from patient files into electronic systems (DAMA and OpenMRS), data cleaning, data quality assurance, and data reporting to regional and national health authorities.

We imputed missing values for orphan status, caregiver’s HIV status, WHO clinical stage of disease, ART regimen at initiation, current ART regimen, and ART dispensation model using multiple imputation by chained equations. We assumed values were missing at random and pooled analyses from 20 sets of imputations using Rubin’s rule ^(21)^. The imputation model included the outcome of retained in care and the following predictors: sex, age, region, urbanicity, age at ART initiation, facility type, facility tier, orphan status, caregiver’s HIV status, WHO clinical stage of disease, ART regimen at initiation, current ART regimen, and ART dispensation model.

### Data analysis

Quantitative data was extracted into an Excel sheet, cleaned, and exported into STATA statistical software version 18.0 (StataCorp, College Station, Texas, USA) for quantitative analysis.

Descriptive statistics were used to characterize the demographic and clinical profiles of children and adolescents at baseline and/or specific time points after initiation of ART. We performed comparisons of retention in care at 6, 12, 18, and 24 months after initiation of ART, comparing sociodemographic and clinical indicators using chi-square tests for proportions. Comparative survival analysis by region was performed, assessing Kaplan-Meier survival estimates. We conducted Cox proportional hazards analyses for the full sample and disaggregated by region using backward stepwise modeling, reporting bivariate and adjusting hazards ratios for retention in care and ART treatment. We included all potential variables in the initial model and removed the least significant one at each step until all remaining variables were statistically significant at the p<0.05 level.

### Ethical Considerations

The study was reviewed and approved by the GU Institutional Review Board in the USA. In Cameroon, ethical review was conducted through the Comité National d’Ethique de la Recherche Pour la Santé Humaine (CNERS) and the Center for Disease Control and Prevention (CDC)-Cameroon. This was a retrospective study of secondary data. All participants were assigned a unique patient code in the database, which did not include any personal patient identifiers. The study team only had access to de-identified secondary data.

## RESULTS

### Demographics, clinical outcomes, and survival estimates at 6, 12, 18, and 24-months after ART initiation

*Table 1* presents the demographic characteristics and clinical history variables of 1,319 ACLHIV enrolled in the ART program across 66 facilities in the Center and East Regions of Cameroon from October 1, 2019, to September 30, 2022. Retention in care rates at the 6-, 12-, 18-, and 24-month intervals were 86.2%, 82.6%, 79.8%, and 77.1%, respectively. *Figure 1* gives the Kaplan-Meier survival estimate for young peoples’ retention in care when enrolled as adolescents in

**Figure 1:**
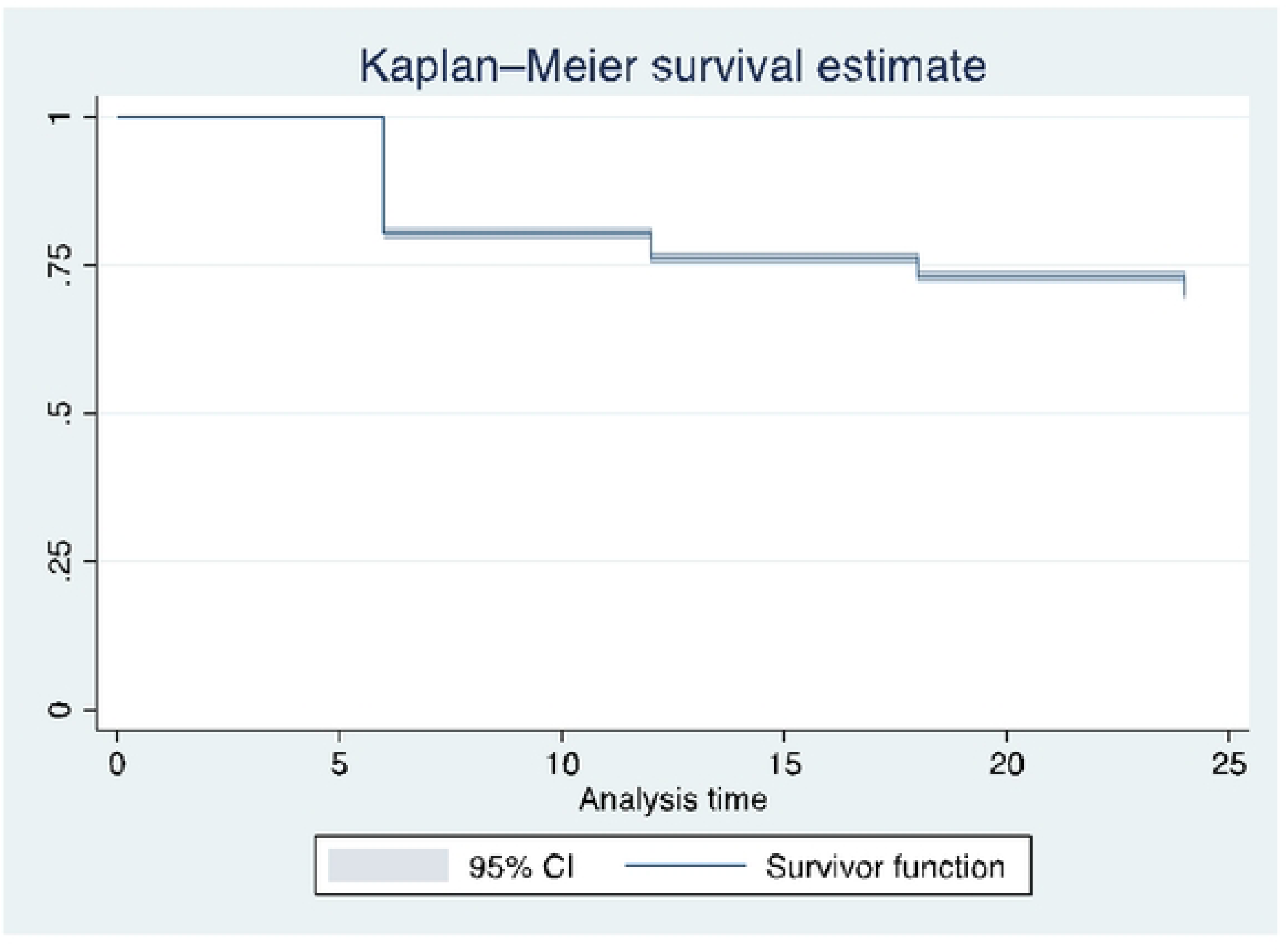
**Kaplan-Meier survival estimates for retention among young people :::,20 years over a two-year period.**

**Table 1:** Retention in care at 6, 12, 18, and 24 months by demographic characteristics and clinical outcomes of 1,319 young people (≤20 years)

|  | Retained in care |  |  |  |  |
| --- | --- | --- | --- | --- | --- |
|  | Total | 6 mo. | 12 mo. | 18 mo. | 24 mo. |
| Overall | % (n=1319) | 86.2 (1137) | 82.6 (1089) | 79.8 (1052) | 77.1 (1017) |
| Region |  |  |  |  |  |
| Center | 48.1 (634) | 89.6 (568) | 87.9 (557) | 86.6 (549) | 83.1 (527) |
| East | 51.9 (685) | 83.1 (569) | 77.7 (532) | 73.4 (503) | 71.5 (490) |
| Urbanicity |  |  |  |  |  |
| Rural | 32.8 (433) | 87.8 (380) | 84.1 (364) | 82.2 (356) | 78.5 (340) |
| Peri/urban | 67.2 (886) | 85.4 (757) | 81.8 (725) | 78.6 (696) | 76.4 (677) |
| Age (in years, current) |  |  |  |  |  |
| 1 – 4 | 28.4 (375) | 81.3 (305) | 73.6 (276) | 70.7 (265) | 68.0 (255) |
| 5 – 9 | 37.2 (491) | 87.2 (428) | 84.9 (417) | 81.1 (398) | 78.4 (385) |
| 10 – 14 | 21.8 (287) | 89.2 (256) | 87.8 (252) | 85.7 (246) | 82.2 (236) |
| 15 – 20 | 12.6 (166) | 89.2 (148) | 86.8 (144) | 86.1 (143) | 84.9 (141) |
| Sex |  |  |  |  |  |
| Male | 49.4 (652) | 85.6 (558) | 81.3 (530) | 78.4 (511) | 75.9 (495) |
| Female | 50.6 (667) | 86.8 (579) | 83.8 (559) | 81.1 (541) | 78.3 (522) |
| Orphan* |  |  |  |  |  |
| Yes | 15.6 (127) | 97.6 (124) | 96.1 (122) | 93.7 (119) | 90.6 (115) |
| No | 84.4 (688) | 90.0 (619) | 86.2 (593) | 83.1 (572) | 81.1 (558) |
| Caregiver HIV status* |  |  |  |  |  |
| Negative | 20.5 (164) | 97.0 (159) | 95.7 (157) | 92.7 (152) | 90.9 (149) |
| Positive | 79.5 (636) | 90.9 (578) | 87.4 (556) | 84.8 (539) | 82.6 (525) |
| Age at ART start (in yrs) |  |  |  |  |  |
| 0 – 4 | 52.8 (696) | 83.1 (578) | 77.4 (539) | 74.0 (515) | 71.7 (499) |
| 5 – 9 | 26.3 (347) | 89.6 (311) | 88.5 (307) | 85.9 (298) | 82.1 (285) |
| 10 – 15 | 20.9 (276) | 89.9 (248) | 88.0 (243) | 86.6 (239) | 84.4 (233) |
| WHO stage* |  |  |  |  |  |
| 1 | 88.3 (1164) | 86.8 (1010) | 83.4 (971) | 80.7 (939) | 78.4 (912) |
| 2 | 4.6 (60) | 86.7 (52) | 83.3 (50) | 78.3 (47) | 76.7 (46) |
| 3 | 6.1 (81) | 81.5 (66) | 75.3 (61) | 72.8 (59) | 65.4 (53) |
| 4 | 1.1 (14) | 64.3 (9) | 50.0 (7) | 50.0 (7) | 42.9 (6) |
| Initial ART* |  |  |  |  |  |
| Non-DTG | 54.9 (645) | 86.1 (555) | 81.2 (524) | 77.2 (498) | 74.9 (483) |
| DTG | 45.1 (530) | 96.8 (513) | 94.9 (503) | 93.2 (494) | 90.0 (477) |
| Current ART* |  |  |  |  |  |
| Non-DTG | 23.3 (275) | 76.4 (210) | 68.7 (189) | 62.9 (173) | 58.6 (161) |
| DTG | 76.7 (903) | 95.7 (864) | 93.5 (844) | 91.7 (828) | 89.3 (806) |
| Dispensation model* |  |  |  |  |  |
| Non-DSD | 47.0 (451) | 74.9 (338) | 68.1 (307) | 63.0 (284) | 60.5 (273) |
| DSD | 53.0 (508) | 96.9 (492) | 95.3 (484) | 93.5 (475) | 91.3 (464) |
| Facility type |  |  |  |  |  |
| Public | 78.2 (1032) | 86.8 (896) | 82.6 (852) | 79.3 (818) | 76.6 (790) |
| Private | 21.8 (287) | 84.0 (241) | 82.6 (237) | 81.5 (234) | 79.1 (227) |
| Facility tier |  |  |  |  |  |
| 1 | 28.7 (379) | 88.4 (335) | 83.1 (315) | 80.2 (304) | 77.8 (295) |
| 2 | 50.8 (670) | 86.1 (577) | 83.0 (556) | 80.6 (540) | 77.5 (519) |
| 3 | 17.5 (231) | 83.1 (192) | 80.1 (185) | 76.6 (177) | 74.9 (173) |
| 4 | 3.0 (39) | 84.6 (33) | 84.6 (4633) | 79.5 (31) | 76.9 (30) |
| * Variable contains some missing data |  |  |  |  |  |

ART within the Center and East Regions. Comparing ACLHIV in each region, those residing in the Center Region were more likely than the East Region to be retained in care at 6 months (89.6% vs. 83.1%), 12 months (87.9% vs. 77.7%), 18 months (86.6% vs. 73.4%), and 24 months (83.1% vs. 71.5%; p=0.000). *Figure 2* compares the Kaplan-Meier survival estimates for retention by region, along with the hazard ratio, showing that ACLHIV in the East Region are 2.66 (CI: 2.51, 2.83; p=0.000) times more likely to fall out of care compared to those in the Center Region. Retention in care increased as adolescents aged at each time point, with those currently between the ages of 1 and 4 years exhibiting the lowest retention at all time points (68.0% at 24 months). There did not appear to be significant differences when comparing urban status and sex; although females were slightly more likely to be retained in care at 6 months (86.8% vs. 85.6%), 12 months (83.8% vs. 81.3%), 18 months (81.1% vs. 78.4%), and 24 months (78.3% vs. 75.9%) compared to males.

**Figure 2:**
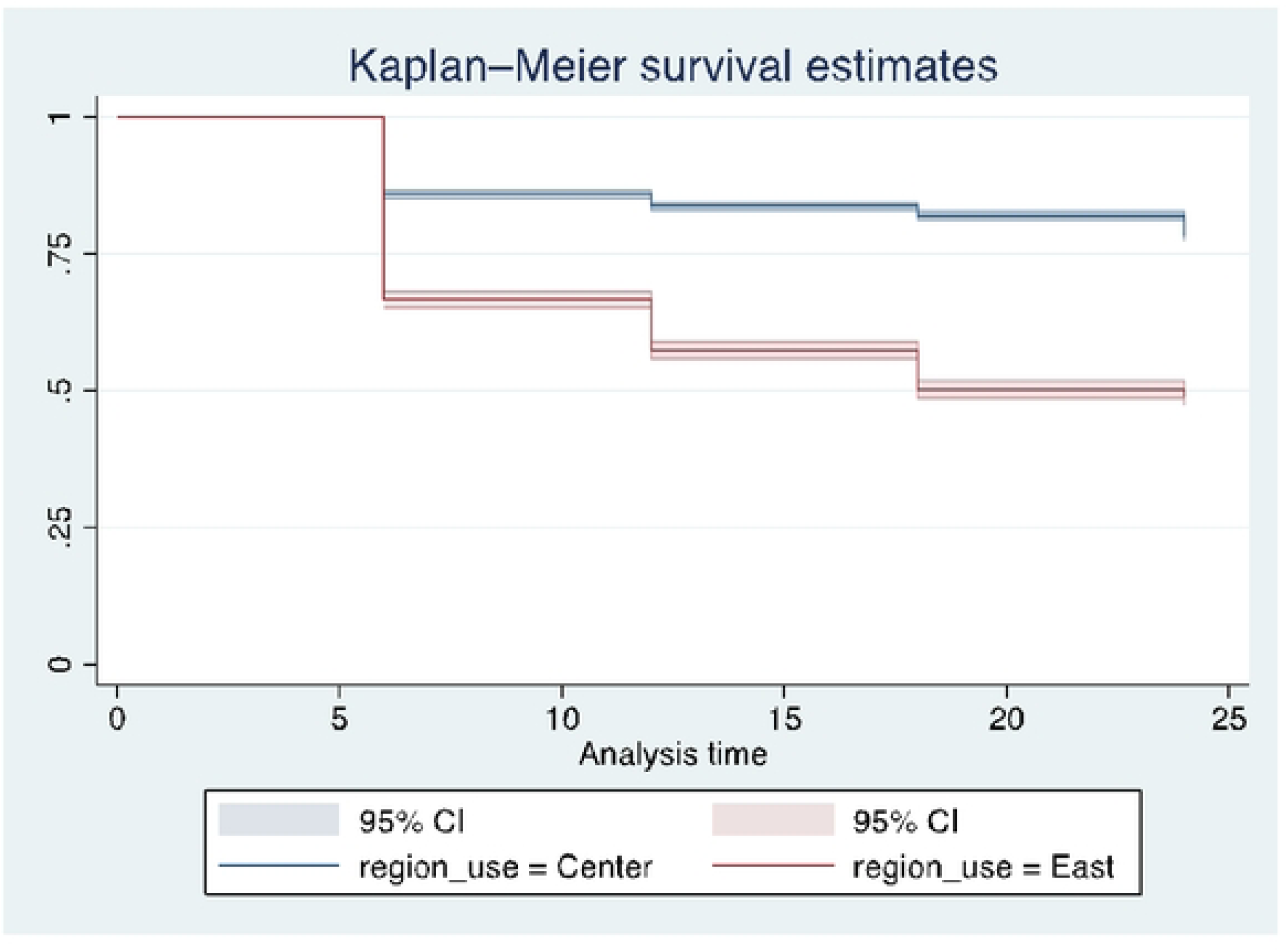
**Kaplan-Meier survival estimates for retention among young people :::,20 years over a two-year period, comparing Center and East Regions, Cameroon.**

Other demographic variables analyzed included orphan status, which appeared to have a protective effect for retention in care. Additionally, the HIV status of the primary caregiver was analyzed, whereby 82.6% of ACLHIV with an HIV+ caregiver were retained at 24 months compared to 90.9% (p=0.10) of ACLHIV with an HIV-caregiver. Assessing clinical outcomes, increasing age at initiation of ART (p=0.000) and lower WHO stages of disease (p=0.001) had higher proportions of ACLHIV retained in care across the four time points. Similarly, taking a DTG-based ART regimen showed higher proportions of ACLHIV retained in care at 6 months (95.7% vs. 76.4%), 12 months (93.5% vs. 68.7%), 18 months (91.7% vs. 62.9%), and 24 months (89.3% vs. 58.6%; p=0.000) compared to those not on a DTG-based ART regimen. ART dispensation also appeared to affect likelihood of retention, with those receiving ART through differentiated service delivery models more likely to be retained in care at each timepoint. At 24 months, 91.3% of ACLHIV receiving ART through a DSD-based model were retained in care, compared to 60.5% (p=0.000) of those receiving ART through a non-DSD model. There were not significant differences across the timepoints for facility type or facility tier.

### Factors associated with retention in care among adolescent living with HIV

*Table 2* presents the results of univariate and multivariate Cox proportional hazards models, adjusted for confounders, for the total sample as well as for each region. In the fully adjusted sample, after applying backward stepwise analysis and multiple imputation for missing values in some predictor variables, the risk of dropping out of care was higher for those residing in peri-/urban areas compared to rural areas (aHR=1.11, 95% CI: 1.03, 1.20) and for ACLHIV with an HIV+ primary caregiver (aHR=1.17, 95% CI: 1.07, 1.28). For WHO stage, the risk decreased from stage 1 to 2, then increased again at stage 3 and 4. Similarly, for age at ART enrollment, the risk was lower for those enrolled at ages 5 – 9 years compared to those older 5 years but increased again for ACLHIV enrolled at ages 10 – 15 years. Facility type did not show a significant association in the adjusted models for the full sample, and regimen at ART initiation was highly correlated with the current ART regimen.

**Table 2:** Cox proportional hazard analysis of factors associated with retention in care over 24 months among adolescents and youth in Center and East Regions, Cameroon.

|  | Total (N=1319) |  | Center Region<br>(n=634) |  | East Region (n=685) |  |
| --- | --- | --- | --- | --- | --- | --- |
|  | crude HR<br>(95% CI) | aHR<br>(95% CI) | crude HR<br>(95% CI) | aHR<br>(95% CI) | crude HR<br>(95% CI) | aHR<br>(95% CI) |
| Urbanicity |  |  |  |  |  |  |
| Rural | 1 | 1 | 1 | -- | 1 | 1 |
| Peri/urban | 1.13 (1.06,<br>1.21) | 1.11 (1.03,<br>1.20) | 1.06 (0.96,<br>1.16) | -- | 0.98 (0.88,<br>1.08) | 1.21 (1.05,<br>1.40) |
| Age (in years, current) |  |  |  |  |  |  |
| 1 – 4 | 1 | 1 | 1 | 1 | 1 | 1 |
| 5 – 9 | 0.66 (0.61,<br>0.71) | 0.66 (0.61,<br>1.16) | 0.60 (0.53,<br>0.67) | 0.57 (0.50,<br>0.64) | 0.84 (0.76,<br>0.93) | 0.82 (0.74,<br>0.92) |
| 10 – 14 | 0.55 (0.51,<br>0.60) | 0.84 (0.71,<br>1.00) | 0.77 (0.69,<br>0.87) | 1.00 (0.78,<br>1.28) | 0.47 (0.40,<br>0.54) | 0.66 (0.56,<br>0.78) |
| 15 – 20 | 0.50 (0.45,<br>0.55) | 0.59 (0.47,<br>0.74) | 0.68 (0.59,<br>0.77) | 0.56 (0.41,<br>0.77) | 0.43 (0.36,<br>0.52) | 0.66 (0.55,<br>0.80) |
| Sex |  |  |  |  |  |  |
| Male | 1 | 1 | 1 | 1 | 1 | 1 |
| Female | 0.87 (0.81,<br>0.92) | 0.88 (0.82,<br>0.94) | 0.71 (0.66,<br>0.78) | 0.74 (0.68,<br>0.81) | 1.04 (0.95,<br>1.13) | 1.12 (1.02,<br>1.22) |
| Orphan* |  |  |  |  |  |  |
| No | 1 | 1 | 1 | 1 | 1 | 1 |
| Yes | 0.44 (0.39,<br>0.49) | 0.72 (0.64,<br>0.82) | 0.57 (0.50,<br>0.65) | 0.76 (0.66,<br>0.89) | 0.33 (0.27,<br>0.40) | 0.61 (0.50,<br>0.74) |
| Caregiver HIV status* |  |  |  |  |  |  |
| Negative | 1 | 1 | 1 | 1 | 1 | -- |
| Positive | 1.66 (1.54,<br>1.80) | 1.17 (1.07,<br>1.28) | 1.26 (1.15,<br>1.38) | 1.16 (1.03,<br>1.29) | 1.63 (1.41,<br>1.89) | -- |
| Age at ART start (in<br>years) |  |  |  |  |  |  |
| 0 – 4 | 1 | 1 | 1 | 1 | 1 | -- |
| 5 – 9 | 0.64 (0.60,<br>0.70) | 0.89 (0.78,<br>1.01) | 0.78 (0.70,<br>0.87) | 0.78 (0.64,<br>0.96) | 0.66 (0.59,<br>0.75) | -- |
| 10 – 15 | 0.57 (0.52,<br>0.62) | 1.04 (0.85,<br>1.28) | 0.81 (0.73,<br>0.90) | 1.07 (0.81,<br>1.41) | 0.44 (0.38,<br>0.50) | -- |
| WHO stage* |  |  |  |  |  |  |
| 1 | 1 | 1 | 1 | 1 | 1 | -- |
| 2 | 0.55 (0.46,<br>0.66) | 0.75 (0.62,<br>0.91) | 0.51 (0.40,<br>0.65) | 0.61 (0.47,<br>0.80) | 0.95 (0.72,<br>1.26) | -- |
| 3 | 1.53 (1.38,<br>1.70) | 1.30 (1.17,<br>1.46) | 1.49 (1.29,<br>1.72) | 1.32 (1.14,<br>1.54) | 1.87 (1.60,<br>2.18) | -- |
| 4 | 2.42 (2.00,<br>2.93) | 1.70 (1.39,<br>2.07) | 3.07 (2.39,<br>3.95) | 2.71 (2.05,<br>3.58) | 1.64 (1.21,<br>2.20) | -- |
| Initial ART* |  |  |  |  |  |  |
| Non-DTG | 1 | -- | 1 | -- | 1 | -- |
| DTG | 0.35 (0.33, 0.38) | -- | 0.36 (0.33, 0.39) | -- | 0.59 (0.53, 0.66) | -- |
| Current ART* |  |  |  |  |  |  |
| Non-DTG | 1 | 1 | 1 | 1 | 1 | 1 |
| DTG | 0.28 (0.27, 0.30) | 0.34 (0.32, 0.36) | 0.26 (0.24, 0.29) | 0.29 (0.27, 0.34) | 0.28 (0.26, 0.31) | 0.35 (0.31, 0.38) |
| Dispensation model* |  |  |  |  |  |  |
| Non-DSD | 1 | 1 | 1 | 1 | 1 | 1 |
| DSD | 0.19 (0.18, 0.21) | 0.23 (0.21, 0.24) | 0.25 (0.23, 0.27) | 0.31 (0.29, 0.34) | 0.15 (0.13, 0.18) | 0.19 (0.16, 0.22) |
| Facility type |  |  |  |  |  |  |
| Public | 1 | -- | 1 | 1 | 1 | 1 |
| Private | 1.00 (0.94, 1.07) | -- | 1.37 (1.26, 1.50) | 1.11 (1.01, 1.21) | 2.34 (2.03, 2.68) | 1.87 (1.57, 2.23) |
| Facility tier |  |  |  |  |  |  |
| 1 | 1 | 1 | 1 | 1 | 1 | 1 |
| 2 | 0.76 (0.71, 0.82) | 1.00 (0.93, 1.09) | 1.39 (1.24, 1.56) | 1.17 (1.03, 1.33) | 0.63 (0.56, 0.70) | 0.99 (0.87, 1.12) |
| 3 | 1.47 (1.34, 1.62) | 1.38 (1.24, 1.53) | 1.57 (1.31, 1.87) | 1.30 (1.07, 1.56) | 1.20 (1.07, 1.34) | 1.31 (1.12, 1.54) |
| 4 | 0.86 (0.74, 1.00) | 1.63 (1.38, 1.93) | 1.08 (0.87, 1.35) | 1.46 (1.17, 1.83) | 2.39 (1.91, 2.99) | 1.92 (1.31, 2.81) |
| * Variable contains some missing data |  |  |  |  |  |  |

Among other factors examined, the risk of falling out of care was lower for females (aHR=0.88, 95% CI: 0.82, 0.94) compared to males. Increasing age was generally associated with a lower risk of dropping out of ART care, with a small rise in risk from ages 5 – 9 years to 10 – 14 years, followed by a reduction in risk for those aged 15 – 20 years compared to those aged 1 – 4 years. Although age was a significant factor, the 5 – 9 and 10 – 14 year age groups were not statistically significantly less likely to drop out of care compared to the 1 – 4 year age group. However, ACLHIV 15 – 20 years had a significantly lower risk of dropping out of care compared to those aged 1 – 4 years. Being an orphan (aHR=0.72, 95% CI: 0.64, 0.82) also decreased the risk of falling out of care compared to ACLHIV with one or more caregivers. Finally, currently taking a DTG-based ART regimen (aHR=0.34, 95% CI: 0.32, 0.36) and receiving ART through DSD-based models (aHR=0.23, 95% CI: 0.21, 0.24) were both associated with a lower risk of falling out of care compared to those on non-DTG and alternative ART delivery models.

*Table 2* also contains adjusted Cox proportional hazards models comparing the Center and East Regions of Cameroon. In the Center Region, the patterns were largely similar to those observed in the total sample. However, urbanicity was not significantly associated with retention in care in the adjusted models, and the regimen at ART initiation remained very highly correlated with the current ART regimen. Additionally, receiving care in a private facility (aHR=1.11, 95% CI: 1.01, 1.21) was associated with an increased risk of falling out of care compared to receiving care at a public facility. The disparity in the risk of falling out of care between young girls and boys living with HIV was more pronounced, with girls exhibiting an even lower risk of dropping out of care compared to the total sample (aHR=0.74, 95% CI: 0.68, 0.81). Similarly, being on a DTG-based ART regimen (aHR=0.29, 95% CI: 0.27, 0.34) and receiving it through DSD-based models (aHR=0.31, 95% CI: 0.29, 0.34) remained associated with a lower risk of falling out of care compared to those on non-DTG regimens and alternative ART delivery models.

In adjusted Cox proportional hazards models for the East Region, several candidate predictors were dropped from the model due to lack of a statistically significant association with retention in care. These predictors include: caregiver HIV status, WHO clinical stage, and age at ART enrollment. Initial ART regimen was highly correlated with current ART and removed from the model. Peri-/urban residence was again associated with a higher risk of falling out of care compared to rural areas. As seen in the Center Region, ACLHIV receiving care at private facilities (aHR=1.87, 95% CI: 1.57, 2.23) were at an increased risk for falling out of care compared to those attending public facilities. Sex patterns shifted in the East Region, with young girls (aHR=1.12, 95% CI: 1.02, 1.22) facing a higher risk of falling out of care compared to young boys. As in the Center Region, being on a DTG-based ART regimen (aHR=0.35, 95% CI: 0.31, 0.38) was associated with a lower risk, with an even stronger protective effect observed for those receiving care through DSD-based models (aHR=0.19, 95% CI: 0.16, 0.22).

## DISCUSSION

This study contributes to the small but growing body of literature on retention in care among youth in SSA ^(5)^. We examined retention rates and associated factors among ACLHIV (0-15 years) receiving ART in the Center and East Regions of Cameroon. Across all time points, retention rates fell below the national and global target of 95%. After 24 months, the ACLHIV retention in the Center Region was 83.1%, and 71.5% in the East Region. These figures document large discrepancies in retention in HIV care for ACLHIV by geographic region in Cameroon. However, they compare favorably to estimates from 2018, which suggested that only 35% of ACLHIV living in Cameroon were effectively on ART treatment ^(23)^. Another study conducted in Cameroon’s East Region found that only 59% of ACLHIV were retained in care after a one-year follow-up period ^(24)^. Examining retention at six-month intervals, lower retention rates emerged early, with 10% of ACLHIV lost-to-follow-up in the Center Region and 17% in the East Region after just six months, considerably higher than losses observed in subsequent six-month periods. Other studies have observed similar ACLHIV retention declines in the first six months ^(25–27)^, underscoring the need for targeted adherence counseling, supportive intervention, and regular follow-up during the early stages of ART initiation for ACLHIV. In addition, access to differentiated ART delivery models is often limited during this high-risk period for disengagement, potentially missing an opportunity to improve early retention ^(15)^.

There are several factors that may help explain the regional disparities in ART care retention among ACLHIV between the Center and East Regions. Prior to the implementation of the TIDE-Cameroon program, in which this study was embedded, the East Region had not benefitted from PEPFAR-funded programming to provide intensive service delivery and patient support. Access to care is particularly challenging in the East Region given the sparse population, long distances between communities and health facilities, and poor road infrastructure. The region hosts displaced populations from the Central Africa Republic, adding further instability and complexity to continued service delivery. Unexpectedly, we observed that ACLHIV located in peri-/urban locations were less likely to be retained in care compared to their rural counterparts, especially in the East Region. While most studies report lower retention among rural ACLHIV due to distance, transport, and infrastructure related challenges ^(12, 28)^, and others have found no effect at all ^(29)^, our findings suggest that urban/peri-urban ACLHIV may face comparable barriers to care, though this cannot be deduced from the study.

Although manifesting in unexpected ways, we found that age categories and sex were associated with ART adherence. First, when compared to older children, we found that ACLHIV children aged less than five were more likely to fall out of care. Meanwhile, older adolescents (>15 years) were the most likely age group to be retained in care. While some comparable studies conducted in SSA found similar results where children aged less than five were most likely to fall out of care ^(30)^. Most studies demonstrated that older adolescents are more likely to fall out of care as they transition from pediatric, to adolescent, to adult HIV care ^(31,32)^. The TIDE program implements a case management system for mothers and their infants, known as the Mother-Baby/Child Case Management System. In this model, client records for the mother and child are linked, and a community health worker is assigned as a case manager to support care navigation and conduct routine check-ins via home or community visits. This intensive monitoring of adherence and continuity of care may help explain some of the retention patterns observed, but our study did not include this variable in our analysis.

Regarding gender disparities, our study found that male ACLHIV were more likely to be retained in care in the Center Region, whereas female ACLHIV were more likely to be retained in the East Region. We hypothesize that this discrepant finding might reflect differing sociopolitical and cultural contexts in the two regions. However, the exact mechanisms by which sex influences retention or non-retention in care among ACLHIV remains unclear. In the literature on ACLHIV in SSA, studies often report conflicting associations between sex and retention in care; some studies show higher retention rates in adolescent boys ^(26,33)^, while others show higher retention rates in adolescent girls ^(34,35)^. Qualitative research suggests that HIV care outcomes are often linked to perceptions of masculinity ^(36)^ and girls taking on caregiving at a young age and/or becoming pregnant ^(37)^.

We anticipated exploring a wider range of caregiver characteristics for associations with retention in ART care and treatment for ACLHIV. However, due to incomplete records, our analysis was limited to examining the HIV status of the primary caregiver, and findings indicated that children with HIV-positive caregivers faced an increased risk of falling out of care. Previous studies have shown that caregiver demographics, treatment outcomes, and caregiver-child relationship dynamics factors are significantly associated with HIV care and treatment outcomes for ACLHIV ^(38–40)^. Nonetheless, caregiver-related factors are often underexplored in studies of ACLHIV retention in care, and there remains a need for high-quality, longitudinal studies to better understand how caregiving influences adolescent adherence in SSA ^(39)^. Additionally, we found that orphaned ACLHIV were more likely to be retained in care compared to ACLHIV with one or more living parents. The literature on orphan status in SSA and retention is mixed, or with null findings ^(41, 42)^. In our study, we hypothesize that the benefits of community-based care models for orphaned ACLHIV in Cameroon potentially outweigh the burden of orphanhood on care retention ^(41)^. Support programs designed for HIV orphaned and vulnerable children may explain this finding, but assessing support from these programs and its impact on retention in HIV care was outside our scope of our study.

Next, we explored select facility characteristics for their influence on ACLHIV retention. First, we found that ACLHIV utilizing private facilities were less likely to be retained in care compared to those using public facilities, particularly in the East Region. Most research on factors associated with ART default in LMIC have focused on government-run health facilities and adult populations, with limited attention given to outcomes in privately-owned facilities. Among those studies which included public and private facilities, Umeokonkwo et al. (2019) ^(43)^ found similar rates of retention for adults in Nigeria, while Chirambo et al. (2019) ^(44)^ found qualitative evidence for higher retention in public facilities in Malawi. These authors linked the choice of public or private facility to factors such as costs, fear of disclosure and stigma, and perceived and actual quality of services. In Cameroon, some authors have discussed the enduring popularity of private and informal health facilities for health and HIV services, citing factors such as those outlined above. Onambele et al. (2023) ^(45)^ further demonstrated that retention in HIV care and treatment is lower among individuals tested in these facilities in urban and semi-urban facilities in Cameroon. We also found that ACLHIV utilizing facilities with higher patient volume were less likely to be retained in care compared to ACLHIV going to lower volume facilities. However, we encountered few studies in SSA that directly explored associations between patient volume and retention in care for ACLHIV, and those that did reported weak or null findings ^(46)^. One study from Kenya found higher retention for HIV-positive infants receiving services from low-volume facilities ^(47)^. In our study, the reasons for these findings remain unclear. The structure and processes for navigating HIV care differ between public and private health facilities. In many settings where HIV services are implemented, community-based support systems are more commonly linked to public health facilities than privately-owned ones, which could partly explain the observed differences in retention. However, our study did not explore this association.

Finally, we explored a range of clinical factors and their association with ACLHIV retention in care. Our analysis found associations with WHO clinical stage of disease, initial and current ART regimen formulation, and ART dispensation models. DSD approaches for ART delivery, including home-based care, family-centered models, CBO programs, PLHIV support groups, or peer-led dispensation, have been implemented at select sites in Cameroon since 2018, with a national framework for DSD scale-up launched in 2022 ^(48)^. DSD models often differ from standard ART delivery by tailoring services for specific populations (e.g., ACLHIV), locations (e.g., community-based dispensing sites), frequencies (e.g., multi-month dispensing), or provider cadres (e.g., community-managed ART delivery) ^(15)^. There is limited, but growing, evidence that DSD approaches to care can improve retention outcomes for ACLHIV in SSA by removing barriers to ART access ^(49, 50)^, but it is still unclear how effective such approaches can be for ACLHIV in supporting retention on ART ^(5)^. In our study, we found clear indications that ACLHIV receiving ART through DSD models were considerably more likely to be retained in care over the two-year follow-up period, suggesting that DSD models could meaningfully expand the reach, effectiveness, and adoption of differentiated care within ACLHIV programs.

We suggest further research to understand which DSD strategies have the greatest effect for ACLHIV in this setting and to better understand the unique access barriers effecting this group.

DTG is the currently recommended first-line ART regimen for LMIC ^(51)^, no longer the lopinavir/ritonavir (LPV/r) regimen. Some studies have found similar or better retention rates for DTG compared to LPV/r ^(51, 52)^, but we are unaware of studies on DTG retention among ACLHIV. In our study, we found that ACLHIV taking DTG as the initial ART-based regimen, as well as those currently on DTG, were considerably more likely to be retained in care compared to those on the LPV/r regimen. Anecdotally, we have some indication that characteristics of the LPV/r drugs (i.e., taste and size) were thought to be unpleasant for ACLHIV. This could account for some of the differences in retention comparing ART formulations, particularly for very young children. Again, we suggest further research to better understand the acceptability of certain characteristics of ART regimens as it relates to our finding that ACLHIV on DTG have better retention outcomes.

This study took place almost entirely within the context of the global COVID-19 pandemic from October 1, 2019, to September 30, 2022. The indirect and potential long-term impacts of the pandemic on food insecurity, lack of medical supplies, loss of income and livelihoods ^(53,54)^, and access to health services at multiple levels were particularly severe in most SSA settings ^(55–57)^. Bachanas et al. (2022) ^(58)^ explored potential disruptions to ART services in Africa at the start of the COVID-19 pandemic, focusing on five countries with the highest HIV burden, including Cameroon. For ACLHIV (< 15 years of age) in Cameroon, they found that the number of children on treatment increased, with a net rise of 42% in new ART initiations, and that the percentage of ACLHIV facing treatment interruptions returned to pre-pandemic levels ^(58)^. The authors suggest that SSA health systems were highly resilient in maintaining ART access and retention among adults, though evidence of similar resilience among ACLHIV populations was more limited ^(58, 59)^. While the Cameroonian health system has seemingly demonstrated resilience in the face of the global pandemic, we caution that the findings from our study should be understood in the context of COVID-19.

### STUDY LIMITATIONS

The primary strength of this study was the large amount of data available for a cohort of ACLHIV over a three-year study period for the analysis, greatly reducing measurement error. Nonetheless, this study had a few limitations. First, this was a retrospective analysis, relying on routinely collected patient data, which was sometimes incomplete. We intended to explore associations among a wider range of sociodemographic, caregiver, and clinical variables, but dropped several due to an unacceptable level of missingness. Missing data from these variables likely introduced non-differential measurement bias. However, we did employ multiple imputation methods for variables which likely avoided some of the potential for such biases. The retrospective nature of the study also subjected it to other risks or confounding factors that may be present but were not measured or included in the model. Second, ACLHIV described as lost-to-follow up may be under-reported due to unrecorded mortality and silent transfers. Silent transfers were treated as still in care, as their records did not list death or loss to follow up under the outcome date and ART outcome.

Another limitation relates to data quality challenges commonly associated with routine program data, though improvements were observed over time. Ongoing data quality assessments and continuous quality improvement initiatives aimed to reduce missing or inaccurate data.

Additionally, as mentioned, the study was conducted during the COVID-19 pandemic, which likely influenced care delivery, decision-making, and retention outcomes for ACLHIV. Despite these limitations, the study sample is likely representative of ACLHIV receiving ART care in two regions of Cameroon, based on the sample size, selection process, and quality control measures.

## CONCLUSION

Nearly one-quarter of ACLHIV enrolled in ART at the beginning of the study period were not retained in care after 24 months. This finding aligns with global trends where adolescents and young people have poorer retention and HIV treatment outcomes compared to adults. ACLHIV remain at high risk of treatment interruption and often loss to follow-up. Our study suggests that retention outcomes in Cameroon are underperforming relative to national ART retention targets.

We identified a range of geographic, sociodemographic, and clinical factors associated with ART retention, all of which should be considered when developing strategies to improve retention in ART programs for ACLHIV. Targeted research, strengthened monitoring and data collection, and enhanced data quality efforts are needed to uncover a wider range of underlying factors, such as caregiver characteristics.

Cameroon, like many LMIC, is characterized by predominantly young population, among whom new HIV infections are rising. Low retention in HIV care among adolescents increases the risk of further HIV transmissions as they reach sexual debut and become sexually active. To achieve and sustain HIV epidemic control, ensuring long-term retention in care for ACLHIV must be a priority. This will require identifying and testing innovative solutions to address societal, health system, contextual, and behavioral factors that undermine treatment adherence and long-term retention in care.

## Data Availability

All relevant data are within the manuscript and its Supporting Information files.

## ACKNOWLEDGEMENTS

This work was supported by the U.S. President’s Emergency Plan for AIDS Relief through the U.S. Centers for Disease Control and Prevention, under Award NU2GGH002214. The views expressed in this manuscript are those of the authors and do not necessarily represent the views of the institutions with which the authors are affiliated.

